# Sex-Specific Associations of Added Sugars and Fructose Exposure Across the First 1,000 Days With Body Composition at 24 Months

**DOI:** 10.64898/2026.10.16.26365755

**Authors:** Sara A. Fortin-Miller, Byron J. Gajewski, Susan E. Carlson, John Colombo, Danielle N. Christifano, Debra K. Sullivan, Holly R. Hull

**Affiliations:** Department of Dietetics and Nutrition, University of Kansas Medical Center, Kansas City, KS; Department of Biostatistics and Data Science, University of Kansas Medical Center, Kansas City, KS; Department of Psychology and Schiefelbusch Institute for Life Span Studies, University of Kansas, Lawrence, KS

**Keywords:** Added sugars, fructose, first 1,000 days, pediatric obesity, body composition, sex differences

## Abstract

**Background:** Added sugars during early development may influence adiposity, but whether associations vary by developmental timing and child sex is unclear.

**Objectives:** To determine whether child sex modifies associations of added sugars and fructose exposure from pregnancy through 24 months with body composition at 24 months.

**Methods:** This secondary longitudinal analysis included 130 mother-child dyads. Maternal diet was assessed during mid-pregnancy and child diet at 2wk and 6mo (infancy), and 12mo and 24mo (toddlerhood). Multivariable regression tested sex-by-exposure interactions for anthropometric outcomes; 42 children had usable dual-energy X-ray absorptiometry (DXA) data.

**Results:** Prenatal added sugars were more positively associated with percentage body fat and total and regional fat mass in girls than in boys. Prenatal fructose was more positively associated with waist circumference in boys. During infancy, added sugars were more positively associated with percentage body fat and total and regional fat mass in boys, whereas fructose was more positively associated with fat-free mass in girls.

**Conclusions:** Associations of early-life sugars exposure with body composition varied by developmental timing and child sex. Findings from the smaller DXA subset are exploratory and require replication but suggest that susceptibility to early dietary sugars may differ across developmental windows.

**Practitioner Points:**

- Repeated dietary assessment from pregnancy through toddlerhood suggested that the relation of early sugars exposure to body composition may differ by both developmental timing and child sex.
- Exploratory DXA findings showed more positive associations of prenatal added sugars with adiposity in girls and of infancy added sugars with adiposity in boys.
- The findings do not support sex-specific dietary recommendations; they reinforce the importance of limiting added sugars in early life and of replicating sex- and timing-specific patterns in larger cohorts.

## Introduction

Nutrition across the first 1,000 days, from conception through 24 months, contributes to growth, body composition, and later metabolic health.^1^ Added sugars (AS) are a modifiable exposure during this period and may displace nutrient-dense foods or contribute to excess energy intake.^2^ Fructose, a major monosaccharide component of AS and a common constituent of sugar-sweetened beverages and processed foods, has been linked to hepatic lipogenesis, insulin resistance, and obesity-related metabolic dysfunction.^3–9^ Defining developmental windows in which sugars exposure is associated with early adiposity may help clarify pathways through which early nutrition contributes to pediatric obesity risk.

Sex differences in growth, fat mass, and fat distribution emerge early in life. Boys generally have greater lean mass accrual and may develop more central fat deposition, whereas girls tend to have a higher proportion of body fat and relatively greater peripheral or subcutaneous fat stores.^10–12^ These divergent trajectories reflect underlying biological differences in adipose tissue development, endocrine signaling, and metabolic programming. However, few studies have examined whether child sex modifies associations between sugars exposure during pregnancy and early childhood and directly assessed body composition. Reliance on weight-based outcomes alone may obscure early differences in fat mass and distribution that are relevant to the development of obesity.

We therefore examined whether child sex modifies associations between AS and fructose exposure across pregnancy, infancy, and toddlerhood and anthropometric and body-composition outcomes at 24 months. We hypothesized that higher sugars exposure during these developmental windows would be associated with greater adiposity at 24 months and that the magnitude and pattern of these associations would differ by child sex.

## Methods

### Study Design and Participants

This secondary analysis included mother-child dyads from two NIH-funded studies: the Assessment of Docosahexaenoic Acid (DHA) on Reducing Early Preterm Birth (ADORE; R01HD83292)^13^ and its offspring follow-up, the Growth and Adiposity in Newborns Study (GAINS; R01DK118220).^14^ ADORE was a multicenter, double-blind, randomized trial in which pregnant individuals (12-20 weeks gestation) received either 1,000 mg/day or 200 mg/day of DHA.^13,15^ Offspring from ADORE were invited to participate in GAINS.^14^ Of the 489 ADORE participants enrolled at the Kansas City site, 254 enrolled their offspring in GAINS. The University of Kansas Medical Center Institutional Review Board approved both studies, and both were conducted in accordance with applicable ethical standards. Before any data collection, participants provided written informed consent for ADORE, and both parents provided written informed consent for their child’s participation in GAINS. ADORE and GAINS were registered at ClinicalTrials.gov (NCT02626299 and NCT03310983).

### Maternal and Offspring Characteristics

Maternal demographics (e.g., race, ethnicity, education, income) and anthropometrics were collected at ADORE enrollment. Offspring demographic characteristics were obtained upon GAINS enrollment.

### Dietary Assessment

Maternal diet at mid-pregnancy (12-20 weeks gestation) was assessed using the NCI Diet History Questionnaire-II (DHQ-II). Data from the DHQ-II were analyzed with Diet*Calc Analysis Software to determine nutrient and food group intake. Because the DHQ-II is not validated for Hispanic populations and may not adequately capture diets that do not reflect a typical US dietary pattern, Spanish-speaking and English-speaking Hispanic participants completed three interviewer-administered multiple-pass 24-hour dietary recalls (two weekdays, one weekend). These recalls were collected by trained research staff fluent in Spanish and analyzed using the Nutrition Data System for Research (NDSR; version 2017, Minneapolis, MN).

Offspring dietary intake was assessed using one interviewer-administered multiple-pass 24-hour dietary recall at 2 weeks, 6 months, 12 months, and 24 months. Energy and nutrient intakes, including AS and fructose, were derived from NDSR. To correct overestimation of AS in infant formulas, where additional lactose is classified as AS, formula-specific adjustment factors provided by NDSR were applied,^16–18^ as previously described.^19^ To represent developmentally distinct postnatal exposure periods, recalls at 2 weeks and 6 months were averaged to represent infancy (Year 1), and recalls at 12 and 24 months were averaged to represent toddlerhood (Year 2); principal component analysis (PCA) with varimax rotation supported this grouping for both AS and fructose. To facilitate interpretation across developmental stages characterized by different energy requirements, AS and fructose were expressed as percent of total energy intake (kcal from sugars ÷ total kcal × 100).

### Anthropometry and Body Composition

At the 24-month visit, trained staff collected anthropometrics using standardized procedures.^14^ Participants were dressed in a clean diaper and weighed on a standing scale; an infant scale was used if a standing measurement could not be collected. Length was measured with a wall-mounted stadiometer; a length board was used if a standing height could not be collected. Weight and length/height were used to calculate weight-for-length (WFL) z-scores. Waist circumference (WC) was measured using a Gulick tape. Skinfold thickness (SKF) was measured at peripheral (biceps, triceps, thigh) and central (suprailiac, subscapular, flank) sites with a Lange caliper. Peripheral and central SKF values were averaged to reflect distribution, consistent with prior methods.^19–21^

Dual-energy X-ray absorptiometry (DXA; GE Lunar Prodigy, encore v13.60) was attempted on all GAINS participants. Specific anatomical landmarks were used to define regions, including arms, legs, and trunk.^14^ To ensure data integrity, a single trained staff member evaluated region demarcation, movement artifacts, and overall scan quality. Consistent with prior literature demonstrating challenges of scanning young children,^21–23^ not all scans were usable; movement artifacts were the primary reason. The DXA analytic subset therefore included children with complete dietary data and a usable DXA scan. Usable total body-less head values were used to derive percentage body fat (%fat), fat mass (FM), fat-free mass (FFM), and regional fat distribution (central and peripheral FM).

### Statistical Analyses

Descriptive characteristics were summarized as mean ± SD or frequencies (%). T-tests and chi-squared tests were used for group comparisons, with Cohen’s d reported for continuous differences. The primary objective was to examine whether sex modified associations between prenatal and early postnatal AS and fructose exposure and body composition at 24 months. This study was a secondary analysis, and sample size was determined by the number of mother-child dyads with complete and usable dietary and outcome data at 24 months.

Hierarchical multivariable linear regression models were constructed separately for anthropometric outcomes (WFL, WC, central SKF, and peripheral SKF) and DXA outcomes (%fat, FM, FFM, central FM, and peripheral FM). Continuous predictors were centered to reduce correlation between interaction terms and their component variables and to improve interpretability of parameter estimates. Full models included:

1. Covariates: maternal age, pre-pregnancy body mass index (BMI), gestational weight gain (GWG), education, smoking status, parity, diagnosis of gestational diabetes (GDM) or a hypertensive disorder of pregnancy (HDP), DHA treatment assignment; offspring race, ethnicity, gestational age, and age at the 24-month visit. Fruit intake was included in fructose models to better distinguish fructose exposure from fruit-related dietary intake.
2. Main effects: child sex (0 male, 1 female), prenatal sugars intake, Year 1 sugars intake, Year 2 sugars intake.
3. Interaction terms: prenatal intake × child sex, Year 1 intake × child sex, Year 2 intake × child sex.

To improve model parsimony given the available sample size, covariates and interaction terms were reduced using backward elimination (removal threshold p>0.10). Primary exposures and child sex were retained in all final models to preserve epidemiologic interpretability. Given the smaller sample size for DXA outcomes (n=42), analyses involving DXA-derived measures were considered exploratory and focused on estimating effect sizes and patterns rather than definitive inference. Model complexity, including interaction terms, was interpreted with caution relative to the available sample size. No adjustment for multiple comparisons was applied given the exploratory nature of these analyses. Statistical significance was defined as p<0.05, with p<0.10 considered approaching significance. All analyses were conducted using IBM SPSS Statistics v29.

## Results

### Sample Characteristics

**Figure 1** depicts participant flow through ADORE and GAINS by treatment assignment. A total of 130 mother-child dyads had complete and usable dietary and anthropometric data. The DXA subset (n=42) was defined as children who had complete dietary data and a usable DXA scan, rather than as a preselected analytic subgroup.

**Figure 1:**
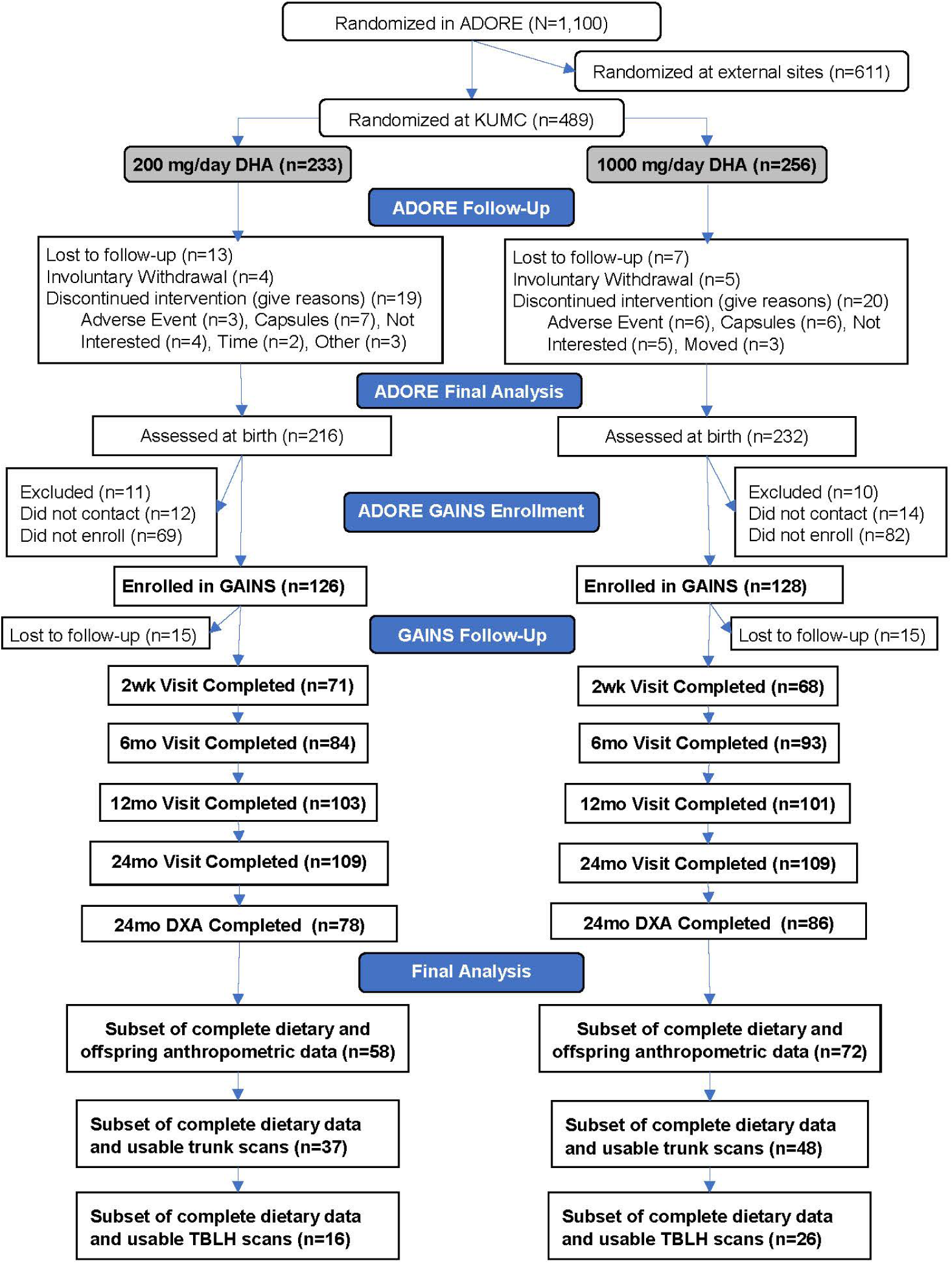
CONSORT diagram of participant flow from the Assessment of Docosahexaenoic Acid (DHA) on Reducing Early Preterm Birth (ADORE) trial into the Growth and Adiposity in Newborns study (GAINS) at the University of Kansas Medical Center (KUMC), showing enrollment, follow-up, and derivation of the final anthropometric cohort and dual-energy x-ray absorptiometry (DXA) subset with complete, usable dietary and outcome data at 24 months of age.

**Table 1** summarizes maternal and offspring demographic characteristics. Mothers were, on average, 31.4 years old with a self-reported pre-pregnancy BMI of 28.2 kg/m² and gained 13.7 kg during pregnancy. Over half (54.6%) exceeded Institute of Medicine GWG guidelines. Most were White (74.6%), non-Hispanic (64.6%), married or partnered (72.3%), parous (60.8%), and had never smoked (76.2%); 85.4% did not develop GDM or HDP. Owing to the Bayesian adaptive design, treatment allocation in ADORE was balanced (55.4% treatment, 44.6% control). Offspring were measured at a mean age of 24.6 months; most were White (66.9%), non-Hispanic (61.5%), and born full-term (92.3%).

**Table 1.**
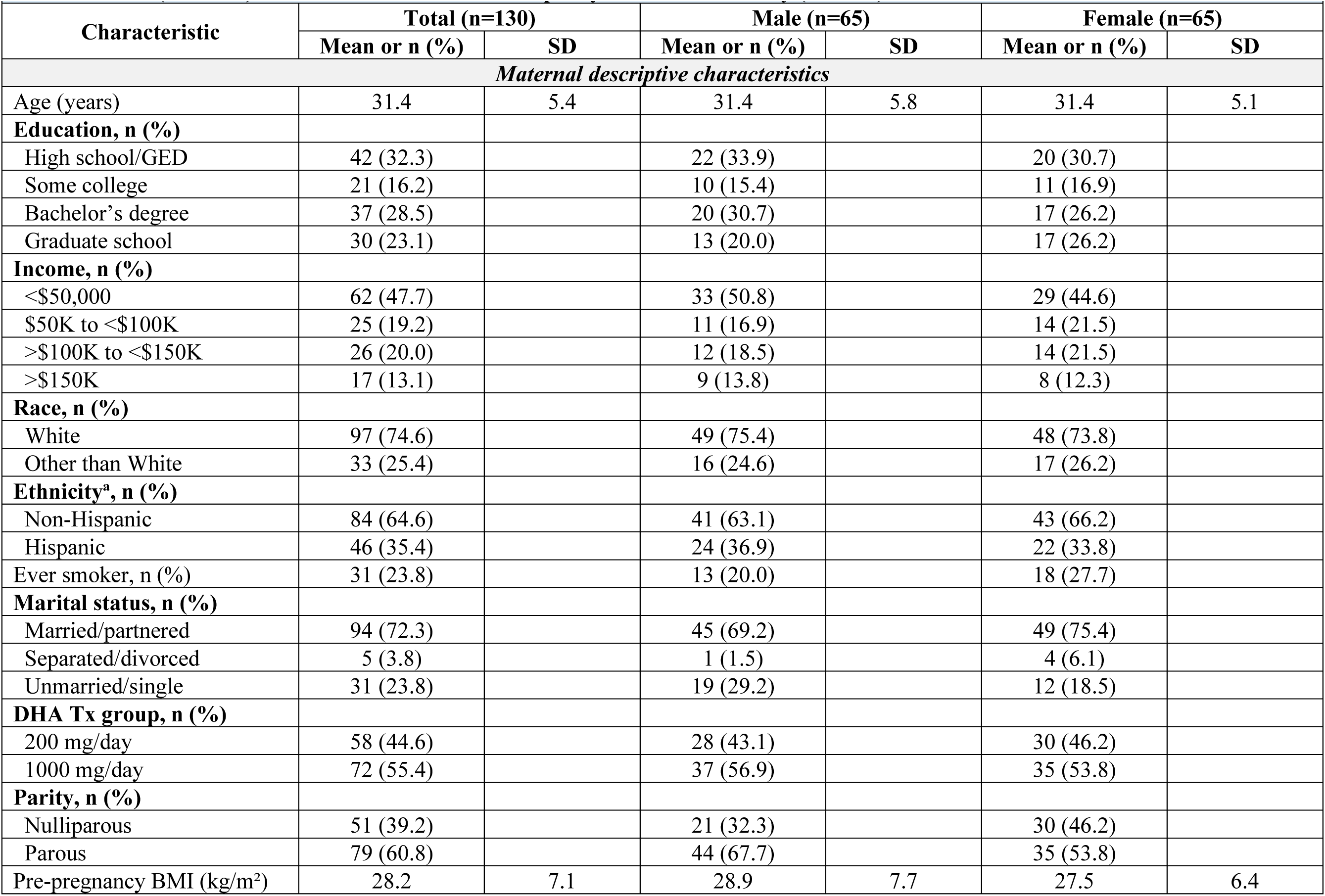
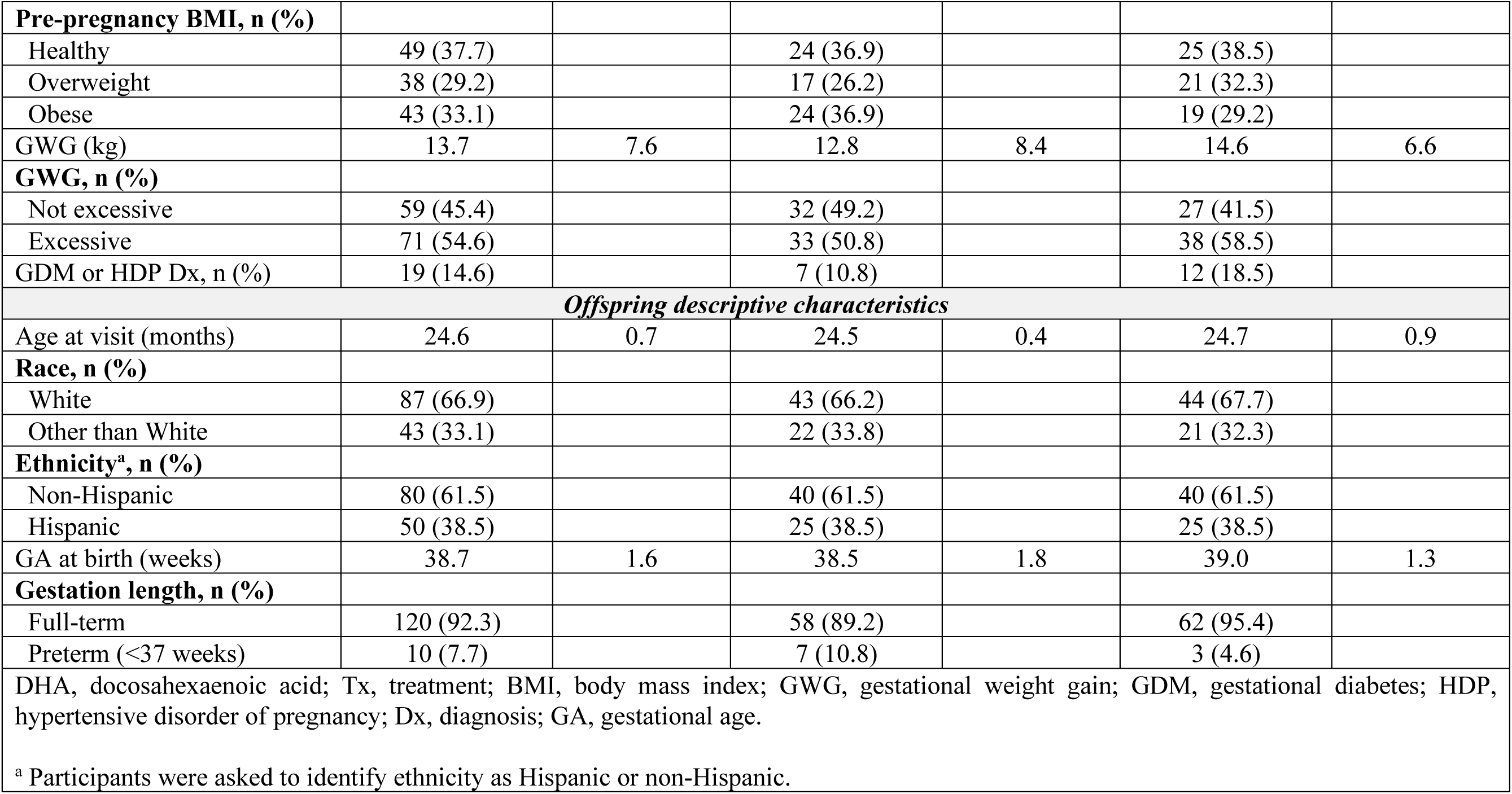
Maternal and offspring descriptive characteristics of mothers and children from the Assessment of DHA on Reducing Early Preterm Birth (ADORE) trial and the Growth and Adiposity in Newborns Study (GAINS)

### Dietary Intake

**Table 2** summarizes maternal and offspring dietary characteristics. Energy and macronutrient intake during pregnancy, Year 1, and Year 2 did not differ by offspring sex. There was a trend toward lower prenatal fruit intake among mothers of female offspring compared with mothers of male offspring (0.6 and 0.8 cup equivalents/1,000 kcal, respectively; p = 0.06). AS and fructose intake were similar between groups (**Figure 2**). Prenatal AS and fructose contributed 12.1% and 5.8% of total energy, respectively. Offspring consumed an average of 2.2% and 6.2% of energy from AS in Year 1 and Year 2, respectively, and 0.7% and 4.6% from fructose in the same periods.

**Table 2.** Maternal and offspring dietary characteristics of mothers and children from the Assessment of DHA on Reducing Early Preterm Birth (ADORE) trial and the Growth and Adiposity in Newborns Study (GAINS)

|  |  |  |  |  |  |  |  |  |
| --- | --- | --- | --- | --- | --- | --- | --- | --- |
| Characteristic | Period | Total (n=130) |  | Male (n=65) |  | Female (n=65) |  | P-value |
|  |  | Mean or n (%) | SD | Mean or n (%) | SD | Mean or n (%) | SD |  |
| Maternal dietary characteristics<sup>a</sup> |  |  |  |  |  |  |  |  |
| Total energy (kcal) |  | 1904.0 | 666.0 | 1842.0 | 647.0 | 1966.0 | 683.0 | 0.29 |
| Carbohydrates (% kcal) |  | 49.3 | 9.2 | 49.2 | 10.0 | 49.4 | 8.4 | 0.91 |
| Fat (% kcal) |  | 35.8 | 7.5 | 35.9 | 8.1 | 35.7 | 6.8 | 0.90 |
| Protein (% kcal) |  | 15.6 | 3.2 | 15.6 | 3.5 | 15.7 | 2.9 | 0.83 |
| AS (g) |  | 61.1 | 59.1 | 58.9 | 61.6 | 63.3 | 57.0 | 0.67 |
| AS (% kcal) |  | 12.1 | 8.7 | 11.8 | 9.2 | 12.4 | 8.3 | 0.68 |
| Fructose (g)<sup>b</sup> |  | 28.5 | 29.6 | 29.2 | 33.7 | 27.8 | 25.2 | 0.79 |
| Fructose (% kcal)<sup>b</sup> |  | 5.8 | 4.5 | 6.0 | 5.3 | 5.6 | 3.7 | 0.66 |
| Fruit servings (cup eq/1000 kcal) |  | 0.7 | 0.6 | 0.8 | 0.7 | 0.6 | 0.4 | 0.06<sup>c</sup> |
| Offspring dietary characteristics<sup>d</sup> |  |  |  |  |  |  |  |  |
| Feeding mode 6mo, n (%) |  |  |  |  |  |  |  |  |
| All/mostly BF |  | 68 (52.3) |  | 37 (56.9) |  | 31 (47.7) |  | 0.57 |
| Equal BF and formula |  | 20 (15.4) |  | 9 (13.8) |  | 11 (16.9) |  |  |
| All/mostly formula |  | 42 (32.3) |  | 19 (29.2) |  | 23 (35.4) |  |  |
| Feeding mode 12mo, n (%) |  |  |  |  |  |  |  |  |
| All/mostly BF |  | 50 (38.5) |  | 27 (41.5) |  | 23 (35.4) |  | 0.76 |
| Equal BF and formula |  | 10 (7.7) |  | 5 (7.7) |  | 5 (7.7) |  |  |
| All/mostly formula |  | 70 (53.8) |  | 34 (52.3) |  | 36 (55.4) |  |  |
| Age of solid food introduction (months) |  | 5.2 | 1.4 | 5.1 | 1.5 | 5.3 | 1.2 | 0.40 |
| Total energy (kcal) | Year 1 | 604.0 | 115.0 | 593.0 | 108.0 | 614.0 | 122.0 | 0.39 |
|  | Year 2 | 1022.0 | 273.0 | 1042.0 | 278.0 | 1001.0 | 269.0 | 0.19 |
| Carbohydrates (% kcal) | Year 1 | 41.6 | 3.5 | 41.5 | 3.3 | 41.7 | 3.7 | 0.80 |
|  | Year 2 | 49.3 | 6.0 | 48.7 | 6.6 | 49.8 | 5.5 | 0.31 |
| Fat (% kcal) | Year 1 | 50.7 | 3.9 | 50.8 | 3.6 | 50.6 | 4.2 | 0.72 |
|  | Year 2 | 36.8 | 5.6 | 36.8 | 6.2 | 36.7 | 4.9 | 0.87 |
| Protein (% kcal) | Year 1 | 7.4 | 1.1 | 7.3 | 1.1 | 7.4 | 1.1 | 0.53 |
|  | Year 2 | 13.8 | 3.2 | 14.3 | 3.5 | 13.4 | 2.8 | 0.10 |
| AS (g) | Year 1 | 3.8 | 8.5 | 4.2 | 9.1 | 3.4 | 7.9 | 0.56 |
|  | Year 2 | 17.2 | 14.0 | 17.0 | 13.0 | 17.4 | 15.0 | 0.86 |
| AS (% kcal) | Year 1 | 2.2 | 4.9 | 2.4 | 5.1 | 2.0 | 4.6 | 0.64 |
|  | Year 2 | 6.2 | 4.2 | 6.0 | 4.1 | 6.4 | 4.4 | 0.55 |
| Any AS intake, n (%) | Year 1 | 48 (36.9) |  | 28 (43.1) |  | 20 (30.8) |  | 0.15 |
|  | Year 2 | 128 (98.5) |  | 64 (98.5) |  | 64 (98.5) |  | 1.00 |
| Fructose (g)<sup>b</sup> | Year 1 | 1.1 | 2.3 | 1.0 | 1.7 | 1.2 | 2.7 | 0.68 |
|  | Year 2 | 11.9 | 7.2 | 12.2 | 7.4 | 11.6 | 7.0 | 0.64 |
| Fructose (% kcal)<sup>b</sup> | Year 1 | 0.7 | 1.5 | 0.6 | 1.0 | 0.7 | 1.8 | 0.66 |
|  | Year 2 | 4.6 | 2.3 | 4.7 | 2.8 | 4.5 | 1.8 | 0.53 |
| Fruit servings (cup eq/1000 kcal) | Year 1 | 0.1 | 0.2 | 0.1 | 0.2 | 0.1 | 0.2 | 0.95 |
|  | Year 2 | 0.8 | 0.6 | 0.8 | 0.6 | 0.8 | 0.5 | 0.78 |
| AS, added sugars; BF, breastfed.<br><sup>a</sup> Mothers' dietary intake was collected between 12 and 20 weeks gestation.<br><sup>b</sup> Fructose is a major monosaccharide component of AS.<br><sup>c</sup> Effect size = 0.33 (small to medium).<br><sup>d</sup> Offspring dietary intake was collected at 2 weeks, 6 months, 12 months, and 24 months. Offspring dietary intake was averaged between 2 weeks and 6 months to represent infancy (Year 1), and between 12 months and 24 months to represent toddlerhood (Year 2). |  |  |  |  |  |  |  |  |

**Figure 2:**
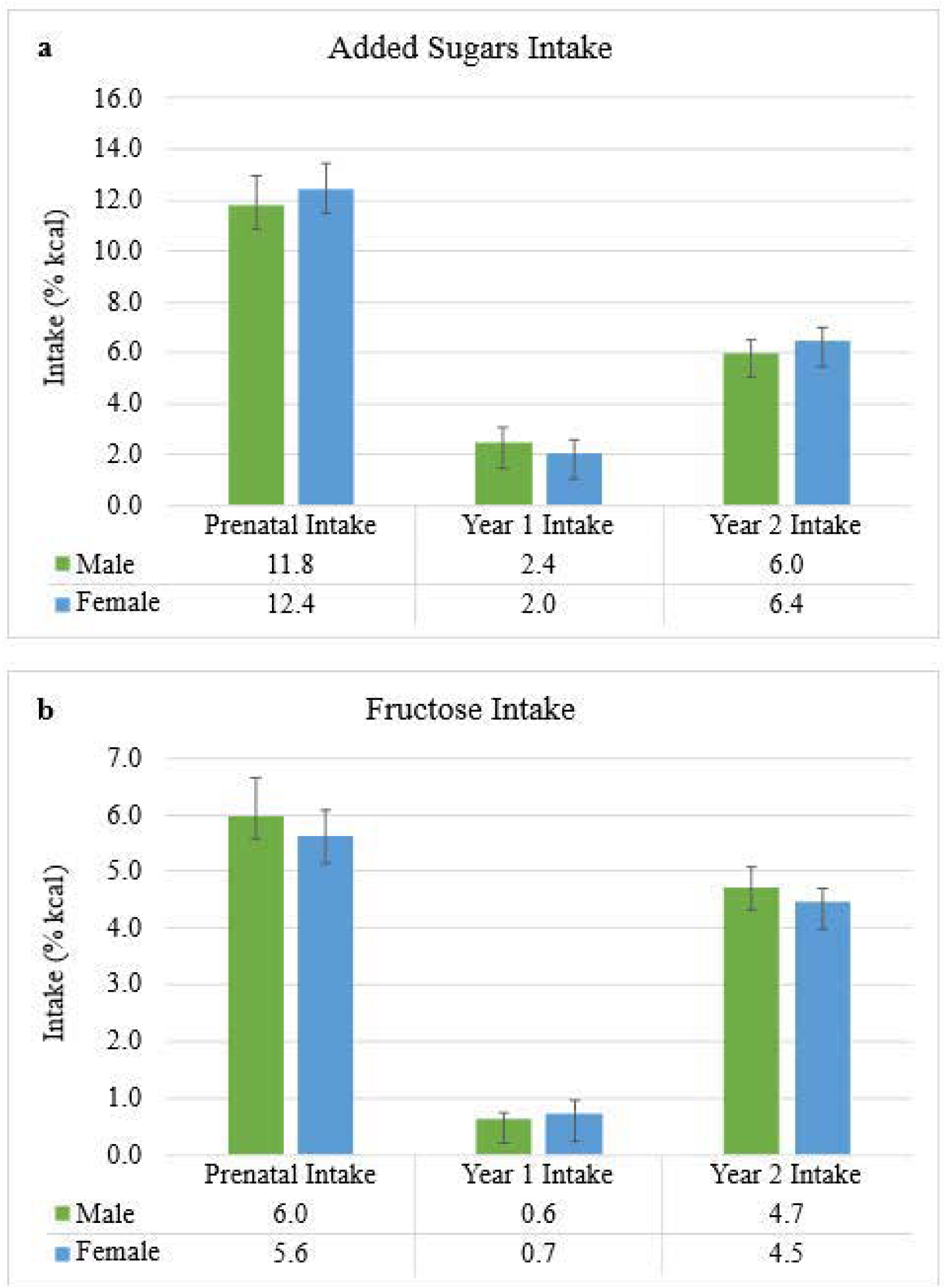
Descriptive comparison of maternal, infant, and toddler sugars intake by offspring sex. Error bars represent means ± SEs. Sugars intake did not differ by offspring sex at any time point. **(a)** Added sugars intake (% of total energy) during pregnancy, infancy (Year 1), and toddlerhood (Year 2). **(b)** Fructose intake (% of total energy) across the same developmental periods.

### Anthropometric and DXA Characteristics

**Table 3** summarizes anthropometric outcomes for the full sample (n=130) and body composition outcomes for the DXA subset (n=42). Anthropometric measures did not differ by sex. In the DXA subset, body composition measures were similar by sex except for FFM, which was lower in females than males (6,894 g vs 7,455 g; p=0.05).

**Table 3.**
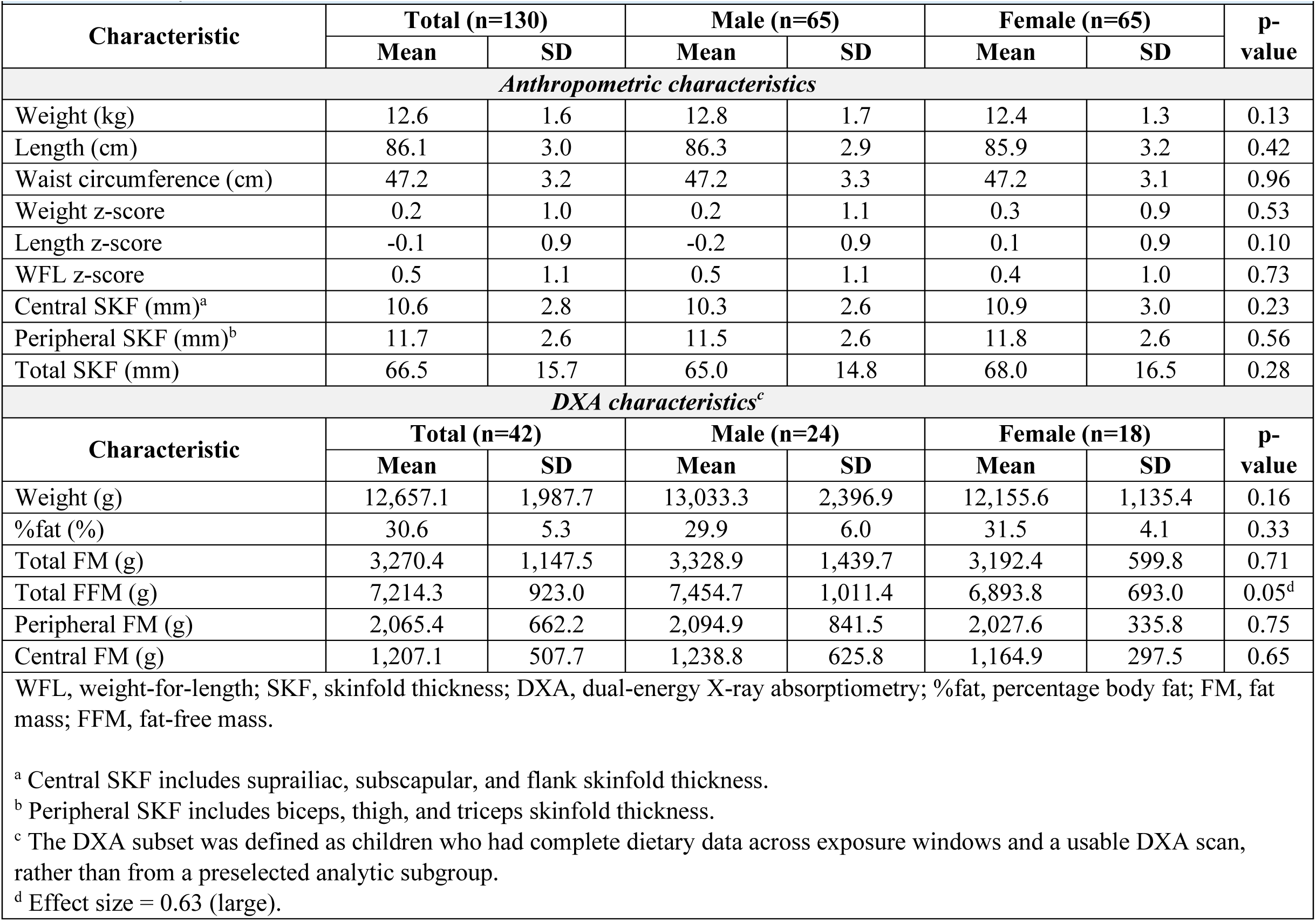
Anthropometric and body composition characteristics of offspring at 24 months in the Growth and Adiposity in Newborns Study (GAINS)

| Characteristic | Total (n=130) |  | Male (n=65) |  | Female (n=65) |  | p-value |
| --- | --- | --- | --- | --- | --- | --- | --- |
|  | Mean | SD | Mean | SD | Mean | SD |  |
| Anthropometric characteristics |  |  |  |  |  |  |  |
| Weight (kg) | 12.6 | 1.6 | 12.8 | 1.7 | 12.4 | 1.3 | 0.13 |
| Length (cm) | 86.1 | 3.0 | 86.3 | 2.9 | 85.9 | 3.2 | 0.42 |
| Waist circumference (cm) | 47.2 | 3.2 | 47.2 | 3.3 | 47.2 | 3.1 | 0.96 |
| Weight z-score | 0.2 | 1.0 | 0.2 | 1.1 | 0.3 | 0.9 | 0.53 |
| Length z-score | -0.1 | 0.9 | -0.2 | 0.9 | 0.1 | 0.9 | 0.10 |
| WFL z-score | 0.5 | 1.1 | 0.5 | 1.1 | 0.4 | 1.0 | 0.73 |
| Central SKF (mm)<sup>a</sup> | 10.6 | 2.8 | 10.3 | 2.6 | 10.9 | 3.0 | 0.23 |
| Peripheral SKF (mm)<sup>b</sup> | 11.7 | 2.6 | 11.5 | 2.6 | 11.8 | 2.6 | 0.56 |
| Total SKF (mm) | 66.5 | 15.7 | 65.0 | 14.8 | 68.0 | 16.5 | 0.28 |
| DXA characteristics<sup>c</sup> |  |  |  |  |  |  |  |
| Characteristic | Total (n=42) |  | Male (n=24) |  | Female (n=18) |  | p-value |
|  | Mean | SD | Mean | SD | Mean | SD |  |
| Weight (g) | 12,657.1 | 1,987.7 | 13,033.3 | 2,396.9 | 12,155.6 | 1,135.4 | 0.16 |
| %fat (%) | 30.6 | 5.3 | 29.9 | 6.0 | 31.5 | 4.1 | 0.33 |
| Total FM (g) | 3,270.4 | 1,147.5 | 3,328.9 | 1,439.7 | 3,192.4 | 599.8 | 0.71 |
| Total FFM (g) | 7,214.3 | 923.0 | 7,454.7 | 1,011.4 | 6,893.8 | 693.0 | 0.05<sup>d</sup> |
| Peripheral FM (g) | 2,065.4 | 662.2 | 2,094.9 | 841.5 | 2,027.6 | 335.8 | 0.75 |
| Central FM (g) | 1,207.1 | 507.7 | 1,238.8 | 625.8 | 1,164.9 | 297.5 | 0.65 |
| WFL, weight-for-length; SKF, skinfold thickness; DXA, dual-energy X-ray absorptiometry; %fat, percentage body fat; FM, fat mass; FFM, fat-free mass. |  |  |  |  |  |  |  |
| <sup>a</sup> Central SKF includes suprailiac, subscapular, and flank skinfold thickness. |  |  |  |  |  |  |  |
| <sup>b</sup> Peripheral SKF includes biceps, thigh, and triceps skinfold thickness. |  |  |  |  |  |  |  |
| <sup>c</sup> The DXA subset was defined as children who had complete dietary data across exposure windows and a usable DXA scan, rather than from a preselected analytic subgroup. |  |  |  |  |  |  |  |
| <sup>d</sup> Effect size = 0.63 (large). |  |  |  |  |  |  |  |

### Associations of Sugars With Anthropometric and DXA Outcomes

Complete main-effects models for anthropometric and DXA outcomes are presented in **Supplementary Tables 1 and 2**, respectively. Results reported below focus on sex-by-exposure interaction terms retained in final adjusted models (**Table 4**), where negative interaction coefficients indicate stronger associations in boys (sex coded 0=male, 1=female).

**Table 4.**
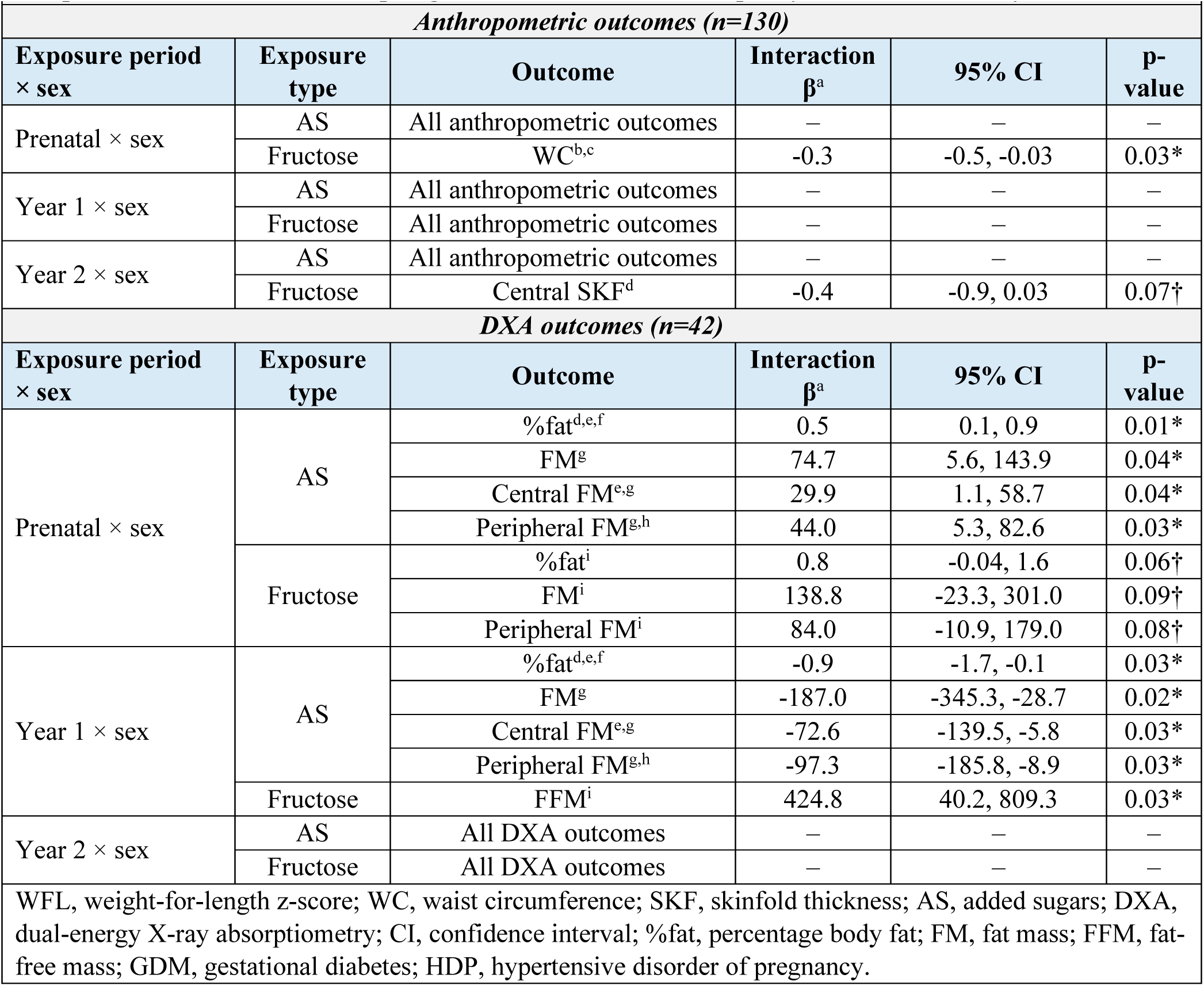
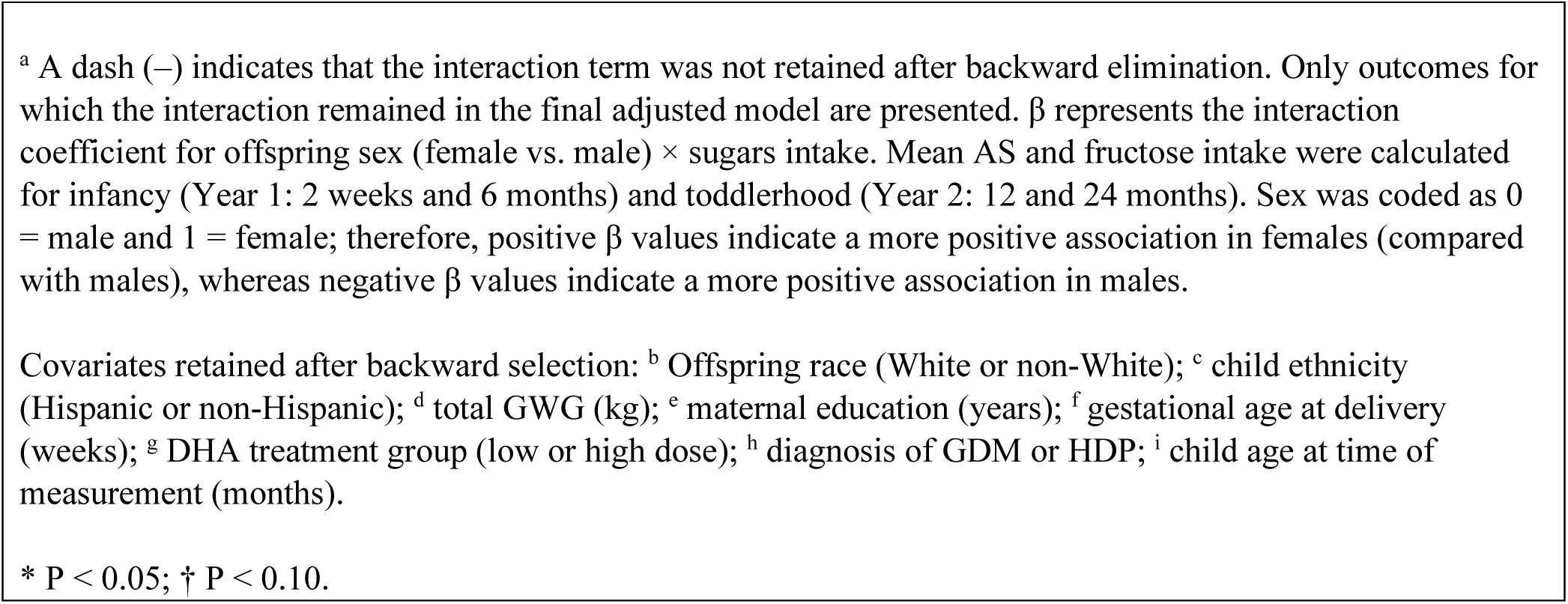
Sex-specific associations between sugars exposure across the first 1,000 days and body composition at 24 months in offspring from the Growth and Adiposity in Newborns Study (GAINS)

|  |  |  |  |  |  |
| --- | --- | --- | --- | --- | --- |
| *Anthropometric outcomes (n=130)* |  |  |  |  |  |
| **Exposure period**<br>**× sex** | **Exposure**<br>**type** | **Outcome** | **Interaction**<br>**$\beta^a$** | **95% CI** | **P-**<br>**value** |
| Prenatal × sex | AS | All anthropometric outcomes | — | — | — |
|  | Fructose | WC<sup>b,c</sup> | -0.3 | -0.5, -0.03 | 0.03* |
| Year 1 × sex | AS | All anthropometric outcomes | — | — | — |
|  | Fructose | All anthropometric outcomes | — | — | — |
| Year 2 × sex | AS | All anthropometric outcomes | — | — | — |
|  | Fructose | Central SKF<sup>d</sup> | -0.4 | -0.9, 0.03 | 0.07† |
| *DXA outcomes (n=42)* |  |  |  |  |  |
| **Exposure period**<br>**× sex** | **Exposure**<br>**type** | **Outcome** | **Interaction**<br>**$\beta^a$** | **95% CI** | **P-**<br>**value** |
| Prenatal × sex | AS | %fat<sup>d,e,f</sup> | 0.5 | 0.1, 0.9 | 0.01* |
|  |  | FM<sup>g</sup> | 74.7 | 5.6, 143.9 | 0.04* |
|  |  | Central FM<sup>e,g</sup> | 29.9 | 1.1, 58.7 | 0.04* |
|  |  | Peripheral FM<sup>g,h</sup> | 44.0 | 5.3, 82.6 | 0.03* |
|  | Fructose | %fat<sup>i</sup> | 0.8 | -0.04, 1.6 | 0.06† |
|  |  | FM<sup>i</sup> | 138.8 | -23.3, 301.0 | 0.09† |
|  |  | Peripheral FM<sup>i</sup> | 84.0 | -10.9, 179.0 | 0.08† |
| Year 1 × sex | AS | %fat<sup>d,e,f</sup> | -0.9 | -1.7, -0.1 | 0.03* |
|  |  | FM<sup>g</sup> | -187.0 | -345.3, -28.7 | 0.02* |
|  |  | Central FM<sup>e,g</sup> | -72.6 | -139.5, -5.8 | 0.03* |
|  |  | Peripheral FM<sup>g,h</sup> | -97.3 | -185.8, -8.9 | 0.03* |
|  | Fructose | FFM<sup>i</sup> | 424.8 | 40.2, 809.3 | 0.03* |
| Year 2 × sex | AS | All DXA outcomes | — | — | — |
|  | Fructose | All DXA outcomes | — | — | — |
| WFL, weight-for-length z-score; WC, waist circumference; SKF, skinfold thickness; AS, added sugars; DXA, dual-energy X-ray absorptiometry; CI, confidence interval; %fat, percentage body fat; FM, fat mass; FFM, fat-free mass; GDM, gestational diabetes; HDP, hypertensive disorder of pregnancy. |  |  |  |  |  |

<sup>a</sup> A dash (–) indicates that the interaction term was not retained after backward elimination. Only outcomes for which the interaction remained in the final adjusted model are presented. $\beta$ represents the interaction coefficient for offspring sex (female vs. male) × sugars intake. Mean AS and fructose intake were calculated for infancy (Year 1: 2 weeks and 6 months) and toddlerhood (Year 2: 12 and 24 months). Sex was coded as 0 = male and 1 = female; therefore, positive $\beta$ values indicate a more positive association in females (compared with males), whereas negative $\beta$ values indicate a more positive association in males.
Covariates retained after backward selection: <sup>b</sup> Offspring race (White or non-White); <sup>c</sup> child ethnicity (Hispanic or non-Hispanic); <sup>d</sup> total GWG (kg); <sup>e</sup> maternal education (years); <sup>f</sup> gestational age at delivery (weeks); <sup>g</sup> DHA treatment group (low or high dose); <sup>h</sup> diagnosis of GDM or HDP; <sup>i</sup> child age at time of measurement (months).
\* $P < 0.05$ ; † $P < 0.10$ .

For anthropometric outcomes, two sex interactions were retained in the final models. Higher prenatal fructose exposure was more positively associated with greater WC in boys than in girls (interaction β= -0.3; 95% CI: (−0.5, -0.03); p=0.03). Year 2 fructose intake approached significance, with a trend toward a more positive association with greater central SKF in boys than in girls (interaction β= -0.4; 95% CI: (−0.9, 0.03); p=0.07). No additional sex interactions were observed for anthropometric outcomes.

Several sex-by-exposure interactions were observed for DXA-derived body composition. Prenatal AS showed the most consistent pattern, with more positive associations in girls than in boys for %fat (interaction β=0.5; 95% CI 0.1, 0.9; p=0.01), total FM (interaction β=74.7; 95% CI 5.6, 143.9; p=0.04), central FM (interaction β=29.9; 95% CI 1.1, 58.7; p=0.04), and peripheral FM (interaction β=44.0; 95% CI 5.3, 82.6; p=0.03). Prenatal fructose showed a less consistent pattern, tending toward more positive associations in girls than in boys for %fat (interaction β=0.8; 95% CI -0.04, 1.6; p=0.06), total FM (interaction β=138.8; 95% CI -23.3, 301.0; p=0.09), and peripheral FM (interaction β=84.0; 95% CI -10.9, 179.0; p=0.08).

Year 1 AS intake was more positively associated with adiposity in boys than in girls, including higher %fat (interaction β= -0.9; 95% CI: (−1.7, -0.1); p=0.03), total FM (interaction β= -187.0; 95% CI: (−345.3, -28.7); p=0.02), central FM (interaction β= -72.6; 95% CI: (−139.5, -5.8); p=0.03), and peripheral FM (interaction β= -97.3; 95% CI: (−185.8, -8.9); p=0.03). Year 1 fructose intake was more positively associated with greater FFM in girls than in boys (interaction β=424.8; 95% CI: (40.2, 809.3); p=0.03). Overall, prenatal AS was more positively associated with adiposity in girls than in boys, whereas postnatal exposures, particularly Year 1 AS and Year 2 fructose, were more positively associated with adiposity-related outcomes in boys than in girls. Prenatal fructose showed sex-specific associations that differed by adiposity measure.

## Discussion

In this longitudinal secondary analysis spanning pregnancy through 24 months, associations between sugars exposure and early body composition differed by developmental timing, sugar type, and child sex. Prenatal AS showed the most consistent pattern of more positive associations with total and regional adiposity in girls than in boys, whereas infancy AS were more positively associated with adiposity in boys. Prenatal fructose was more positively associated with WC in boys, and infancy fructose was more positively associated with FFM in girls. These patterns suggest that developmental timing and biological sex may jointly influence susceptibility to early dietary exposures. Because the DXA analyses were based on a smaller subset, those findings should be considered exploratory and hypothesis-generating rather than definitive evidence of sex-specific effects.

### Developmental Timing and Sex-Specific Susceptibility

The observed patterns are biologically plausible in the context of established sex differences in early growth and body composition. Females generally have a higher proportion of body fat, whereas males accrue more lean mass, grow more rapidly early in postnatal life, and tend toward greater central fat deposition.^11,24–26^ These trajectories begin early and may create different responses to nutritional exposures across development.^11,27^ In the present study, prenatal AS were more positively associated with %fat and total, central, and peripheral FM in girls, suggesting greater susceptibility of overall adiposity to the prenatal nutritional environment. Prenatal fructose showed a less consistent pattern, including a more positive association with WC in boys.

Sexual dimorphism in placental function, fetal endocrine signaling, and developmental plasticity may contribute to these differences. Experimental and observational work suggests female fetuses show greater developmental plasticity in response to maternal nutritional environments, promoting greater overall fat accretion under conditions of higher energy or sugar availability.^28–30^ In contrast, male fetuses may be less globally plastic but more vulnerable to perturbations in pathways governing fat distribution.^29,31^ Such mechanisms could help explain why prenatal AS tracked more strongly with overall adiposity in girls while prenatal fructose was associated with a marker of central adiposity in boys. These mechanistic interpretations remain speculative in the absence of placental, endocrine, or metabolic biomarkers and should guide future studies rather than be viewed as causal explanations of the present findings.

Postnatally, the pattern shifted toward greater adiposity-related susceptibility in boys. During infancy, AS were more positively associated with %fat and total and regional FM in boys than in girls, whereas fructose was more positively associated with FFM in girls. Infancy is characterized by rapid somatic growth and high anabolic demands,^25,32^ and sex differences in energy partitioning may contribute to divergent associations with fat and lean tissue, particularly in boys who accrue lean mass rapidly. By toddlerhood, fructose intake approached significance for a more positive association with central adiposity in boys. As growth velocity slows in toddlerhood and regulation of energy partitioning increases, excess sugars may be more readily stored as fat. In this context, increasing fructose exposure as diets diversify during toddlerhood could contribute to greater central adiposity among boys, potentially amplifying typical sex differences in fat distribution.^27^ Together, these findings support examining pregnancy, infancy, and toddlerhood as distinct developmental windows rather than treating early-life diet as a single exposure period.

### Sugar Type and Body-Composition Phenotype

AS showed more consistent associations with total and regional adiposity, whereas fructose was associated with selected measures of fat distribution and FFM. This distinction may reflect both biological and measurement differences. Total AS capture a broader dietary exposure, while fructose has distinct hepatic metabolic pathways linked to *de novo* lipogenesis and central fat accumulation.^3–5,33,34^ At the same time, fructose intake in early life arises from heterogeneous food and beverage sources, including naturally occurring sources, and the present observational data cannot isolate specific metabolic mechanisms. Direct measures of fat and lean mass therefore add useful phenotypic resolution but do not establish causal pathways.

### Implications for Pediatric Obesity Prevention

Current U.S. dietary guidance emphasizes limiting AS across childhood; guidance for infants and toddlers specifically recommends avoiding AS before age 2.^35,36^ The present findings do not support different intake recommendations for boys and girls. Rather, they suggest that the relation of early dietary sugars to emerging adiposity may vary across developmental windows and that studies of pediatric obesity risk should consider both timing of exposure and sex. From a prevention perspective, pregnancy and the first two years remain important periods for establishing dietary patterns and limiting unnecessary AS, while larger prospective studies are needed to determine whether reducing exposure alters subsequent body-composition trajectories.

## Strengths and Limitations

Strengths include the longitudinal assessment of sugars exposure across pregnancy, infancy, and toddlerhood; use of validated dietary assessment methods; and complementary measures of growth and adiposity. WFL is an important marker of early growth but does not distinguish fat from lean tissue. The inclusion of WC and SKF in the full cohort, together with DXA-derived total and regional fat and lean mass in a subset, provides more direct and compartment-specific characterization of body composition.^37,38^ Another strength was refinement of infant AS estimates by adjusting NDSR output to distinguish lactose added to infant formula from true AS, reducing exposure misclassification during a period when formula may constitute a large proportion of energy intake.

Several limitations are important. Only one offspring 24-hour dietary recall was collected at each time point, which may have increased measurement error relative to repeated recalls for estimating usual intake.^39^ PCA supported grouping the postnatal assessments into infancy and toddlerhood, but these averages may still obscure within-period variation. Breastmilk composition was not directly measured; standardized volumes and NDSR-derived nutrient estimates were used instead.^40–42^ Usable DXA data were available for 42 children, largely because motion-free scans are difficult to obtain at this age. The smaller subset reduced precision for interaction estimates and limits generalizability of the body-composition findings. Finally, this secondary analysis was not powered *a priori* for sex-by-exposure interactions, included multiple outcomes and developmental windows, and did not adjust for multiple comparisons. Accordingly, the interaction findings, particularly those from DXA, should be interpreted as exploratory patterns requiring replication in larger cohorts.

## Future Directions

Future studies should replicate these associations in larger and more diverse cohorts using repeated dietary recalls and repeated body-composition measurements across childhood. Priority areas include identifying the food and beverage sources of AS and fructose, determining whether specific sources are differentially associated with adiposity, and integrating placental, endocrine, metabolic, and food-environment measures that may clarify sex-specific mechanisms. Longitudinal follow-up is also needed to determine whether the early differences observed here persist, attenuate, or predict later obesity and cardiometabolic risk.

## Conclusion

Across the first 1,000 days, associations of AS and fructose exposure with body composition at 24 months differed by developmental timing and child sex. Prenatal AS were more positively associated with adiposity in girls, whereas infancy AS were more positively associated with adiposity in boys. These findings do not establish causal or sex-specific dietary effects, but they highlight developmental timing and sex as potentially important sources of heterogeneity in early obesity risk. Replication in larger cohorts with repeated dietary and body-composition measures is needed to determine the robustness and longer-term significance of these patterns.

## Acknowledgments

The authors would like to acknowledge and thank the participants involved in the study. They would also like to thank current and previous members of the Hull Laboratory that assisted in data collection.

## Author contributions

S.A.F.M. conducted the research, analyzed data, performed statistical analyses, and drafted the manuscript. B.J.G. contributed to study design and statistical analysis.

S.E.C. contributed to study design and manuscript development. J.C. contributed to statistical analysis. D.N.C. contributed to manuscript development. D.K.S. provided essential materials. H.R.H. contributed to study design and manuscript development and had primary responsibility for final content. All authors reviewed and approved the final manuscript.

## Data availability

Data described in the manuscript will be made available upon reasonable request.

## AI disclosure

During manuscript preparation, the authors used OpenAI ChatGPT to assist with language editing, clarity, and organization. The authors reviewed and edited all generated content and take full responsibility for the accuracy, integrity, and final content of the manuscript.

