## Supplementary Tables 1 and 2 for "Sex-Specific Associations of Added Sugars and Fructose Exposure Across the First 1,000 Days With Body Composition at 24 Months"

### Supplementary Materials

#### Supplementary Results

Main-effects associations between AS and fructose exposure across pregnancy, infancy (Year 1), and toddlerhood (Year 2) and offspring anthropometric outcomes at 24 months are presented in **Supplementary Table 1**. Higher AS intake during Year 1 was associated with higher WFL z-score and peripheral SKF, with a trend toward higher central SKF. Year 2 AS intake showed a trend toward lower WFL z-score. For fructose, Year 1 intake was associated with higher WFL z-score and WC and showed a trend toward higher central SKF.

Main-effects associations with DXA-derived body composition outcomes among dyads with complete dietary data and usable DXA scans are shown in **Supplementary Table 2**. Higher AS intake during Year 1 was associated with greater FFM, whereas higher AS intake during Year 2 was associated with lower %fat, total FM, and both central and peripheral FM. In contrast, higher fructose intake during Year 2 was positively associated with %fat, total FM, and central and peripheral FM, with a trend toward greater FFM. Sex × exposure interaction terms were tested in all models and removed when not retained following backward elimination; retained interaction effects are presented in the primary manuscript. Covariates retained in final adjusted models varied by outcome and exposure window and are summarized in each table.

#### Supplementary Discussion

The main-effects associations observed across the first 1,000 days are consistent with prior literature linking early-life sugars exposure to variation in growth and adiposity during early childhood. Associations differed by developmental window and outcome, highlighting the importance of exposure timing when evaluating early dietary influences on growth. Interpretation of these findings is informed by the sex-specific interaction effects emphasized in the primary manuscript, which suggest differential susceptibility to early sugars exposure across developmental periods.

**Supplementary Table 1. Main-effects associations between added sugars and fructose exposure across the first 1,000 days and offspring anthropometric outcomes at 24 months (n=130)**

| Independent variable | WFL (z-score) | | WC (cm)[a,d,e] | | Central SKF (mm)[b] | | Peripheral SKF (mm)[c,f] | |
|---|---|---|---|---|---|---|---|---|
| | β | P | β | P | β | P | β | P |
| ***Added sugars*** | | | | | | | | |
| Offspring sex | -0.03 | 0.89 | 0.26 | 0.64 | 0.52 | 0.29 | 0.33 | 0.47 |
| Prenatal intake | 0.01 | 0.83 | -0.05 | 0.19 | 0.01 | 0.79 | 0.01 | 0.70 |
| Year 1 intake | 0.05 | 0.01* | 0.01 | 0.16 | 0.09 | 0.10 | 0.14 | 0.01* |
| Year 2 intake | -0.04 | 0.06† | -0.11 | 0.13 | -0.03 | 0.63 | -0.08 | 0.14 |
| ***Fructose*** | | | | | | | | |
| Offspring sex | -0.06 | 0.74 | 1.49 | 0.10 | 2.35 | 0.04* | 0.07 | 0.88 |
| Prenatal intake | 0.01 | 0.79 | 0.12 | 0.13 | 0.02 | 0.68 | 0.02 | 0.65 |
| Year 1 intake | 0.13 | 0.04* | 0.46 | 0.02* | 0.29 | 0.08† | 0.26 | 0.11 |
| Year 2 intake | 0.01 | 0.71 | 0.17 | 0.14 | 0.01 | 0.94 | -0.07 | 0.46 |

AS, added sugars; BMI, body mass index; GWG, gestational weight gain; SKF, skinfold thickness; WC, waist circumference; WFL, weight-for-length; Year 1, infancy (2 weeks and 6 months); Year 2, toddlerhood (12 and 24 months).

[a] Added sugars/WC model retained pre-pregnancy BMI.
[b] Added sugars/central SKF and fructose/central SKF models retained total GWG (kg).
[c] Added sugars/peripheral SKF model retained maternal education (years).
[d] Fructose/WC model retained offspring race (White or non-White).
[e] Fructose/WC model retained child ethnicity (Hispanic or non-Hispanic).
[f] Fructose/peripheral SKF model retained parity (nulliparous or parous).

* $P < 0.05$; † $P < 0.10$.

**Supplementary Table 2. Main-effects associations between added sugars and fructose exposure across the first 1,000 days and offspring DXA-derived body composition outcomes at 24 months (n=42)**

| Independent variable | %fat (%)[a,b,c,h] | | Total FM (g)[d,h] | | Total FFM (g)[d,e,f,h] | | Central FM (g)[a,b,d,h,i] | | Peripheral FM (g)[d,g,h] | |
|---|---|---|---|---|---|---|---|---|---|---|
| | β | P | β | P | β | P | β | P | β | P |
| ***Added sugars*** | | | | | | | | | | |
| Offspring sex | 1.41 | 0.40 | -387.22 | 0.20 | -609.67 | 0.01* | -154.27 | 0.22 | -202.21 | 0.23 |
| Prenatal intake | -0.35 | 0.05† | -45.24 | 0.10 | 7.18 | 0.68 | -21.93 | 0.06† | -24.46 | 0.12 |
| Year 1 intake | 0.24 | 0.29 | 150.37 | <0.01* | 101.68 | <0.01* | 55.03 | 0.01* | 89.72 | <0.01* |
| Year 2 intake | -0.61 | 0.02* | -105.78 | 0.02* | -42.91 | 0.21 | -46.40 | 0.01* | -66.09 | 0.01* |
| ***Fructose*** | | | | | | | | | | |
| Offspring sex | -0.81 | 0.76 | -423.49 | 0.44 | -485.72 | 0.09† | 35.86 | 0.82 | -252.41 | 0.43 |
| Prenatal intake | -0.52 | 0.05† | -95.85 | 0.07† | -26.82 | 0.38 | 4.17 | 0.82 | -58.97 | 0.06† |
| Year 1 intake | -0.70 | 0.26 | -105.23 | 0.40 | -215.18 | 0.12 | -46.59 | 0.42 | -73.77 | 0.32 |
| Year 2 intake | 0.91 | 0.03* | 212.79 | 0.01* | 102.18 | 0.10 | 73.76 | 0.05† | 115.88 | 0.02* |

AS, added sugars; BMI, body mass index; DHA, docosahexaenoic acid; DXA, dual-energy X-ray absorptiometry; FFM, fat-free mass; FM, fat mass; GDM, gestational diabetes; GWG, gestational weight gain; HDP, hypertensive disorder of pregnancy; %fat, percentage body fat; Year 1, infancy (2 weeks and 6 months); Year 2, toddlerhood (12 and 24 months).

[a] Total GWG (kg) was retained in added sugars/%fat and fructose/central FM models.
[b] Maternal education (years) was retained in added sugars/%fat and added sugars/central FM models.
[c] Gestational age at delivery (weeks) was retained in the added sugars/%fat model.
[d] DHA treatment group (low or high dose) was retained in added sugars/FM, FFM, central FM, and peripheral FM models.
[e] Pre-pregnancy BMI was retained in the added sugars/FFM model.
[f] Ever smoker (yes/no) was retained in the added sugars/FFM model.
[g] Diagnosis of GDM or HDP was retained in the added sugars/peripheral FM model.
[h] Child age at time of measurement (months) was retained in all fructose models.
[i] Child ethnicity (Hispanic or non-Hispanic) was retained in the fructose/central FM model.

* $P < 0.05$; † $P < 0.10$.
